# Corticospinal tract risk modifies motor recovery after minimally invasive surgery for intracerebral hemorrhage: a secondary analysis of MISTIE-III

**DOI:** 10.64898/2026.06.10.26354920

**Authors:** Olivia N Murray, David A Jenkins, Nathan Walborn, Hiren C Patel, George Harston, Timothy Cootes, Catharina JM Klijn, Wendy Ziai, Daniel F Hanley, Ulrike Hammerbeck, Adrian R Parry-Jones

## Abstract

**Objective:** Outcome after surgical hematoma evacuation for intracerebral hemorrhage (ICH) depends on hematoma location. As corticospinal tract (CST) integrity affects motor recovery after stroke, we hypothesized that CST integrity drives heterogeneity in surgical outcomes and investigated this in a secondary analysis of MISTIE-III participants.

**Methods:** Risk of CST injury was categorized into four levels, based on the interaction between the CST, the hematoma, and perihematomal edema (PHE) on automatically segmented stability CT: no risk, PHE infiltration, hematoma infiltration, and complete interruption of the CST. Associations with outcome were tested using multivariable linear regression for motor National Institutes of Health Stroke Scale (NIHSS) at day 180 and ordinal regression for modified Rankin Scale (mRS) at day 365, introducing an interaction term between CST risk and treatment group.

**Results:** Day 180 motor NIHSS was significantly lower for ‘no risk’ (b:-3.77, [95% confidence interval [CI]: -5.8 to -1.70], *p*=0.0003) and ‘PHE infiltration’ (b:-2.3, [95%CI: - 3.5 to -1.1]; *p*=0.0002) vs. ‘complete interruption’. Surgery was associated with lower Day 180 motor NIHSS in participants with hematoma infiltration (b:-2.07, [95%CI: -3.8 to -0.4], *p*=0.016). Compared to complete interruption, ‘no risk’ (adjusted odds ratio [aOR]:0.27, [95%CI: 0.10 to 0.74], *p*=0.01) and ‘PHE infiltration’ (aOR:0.41, [95%CI: 0.23 to 0.74]; *p*=0.003) were associated with lower odds of unfavorable day 365 mRS. Surgery was associated with lower mRS in participants with no risk (aOR:0.23, [95%CI: 0.05 to 0.97, *p*=0.045).

**Interpretation:** Increasing CST risk is associated with worse motor recovery (day 180) and disability (day 365). CST risk modifies the effect of the MISTIE-III procedure on motor recovery and disability.

**Trial registration:** NCT01827046 (https://clinicaltrials.gov/study/NCT01827046)

## Introduction

Intracerebral hemorrhage (ICH) is a devastating form of stroke. One third of all ICH patients die in hospital, and survivors are often left with significant disability.^1^ Despite recent advances in care, there are limited effective treatment options.^2^ A growing evidence base suggests that minimally invasive surgical (MIS) hematoma evacuation may reduce mortality and improve functional outcome^3^. The MISTIE-III trial (n=499) investigated MIS with thrombolysis and did not demonstrate a significant improvement in functional outcomes.^4^ Subsequently, the ENRICH trial (n=300) tested MIS within 24 hours of ictus in patients with ICH volume between 30 and 80 ml and showed improved functional outcome, unless ICH was in the anterior basal ganglia (31% of participants)^5^. The recent MIND trial (n=236) tested MIS within 72 hours of ictus in patients with ICH volume between 20 and 80 ml^6^. MIND was neutral for mRS at day 180 but recruited a higher proportion of deep ICHs (70%). These trials suggest that location may be a key factor in whether MIS improves functional outcome.

The functional and structural integrity of the corticospinal tract (CST) is known to influence the trajectory of motor recovery after both ischemic stroke and ICH,^7–11^ with the extent of injury determining the magnitude of functional impairment.^12^ ICH may completely and irreversibly disrupt the CST^13^ or it may infiltrate only a proportion of tract fibers. Further, if the ICH is adjacent to but not in contact with the CST, secondary injury may occur, associated with perihematomal edema (PHE).^14^ Functional integrity of the CST is typically measured using transcranial magnetic stimulation,^11^ and structural integrity can be measured using diffusion tensor MRI.^10^ However, both require specialist equipment and analysis and are not routinely available in clinical settings^15^. We have previously developed a deep learning method for estimating CST location on non-contrast diagnostic ICH CT scans and shown that the extent to which the ICH overlaps with the predicted CST location is strongly associated with motor outcome^16,17^.

Here, we developed a four-level, automatically computed assessment of CST injury risk and applied it to a secondary analysis of MISTIE-III trial participants. We hypothesized that MIS may improve outcome when the CST is at risk of injury from hematoma infiltration and/or secondary injury and be of no benefit where irreparable damage has occurred by complete CST disruption by the ICH.

## Methods

### Participants

This study is a retrospective, secondary analysis of the Minimally Invasive Surgery plus Thrombolysis for Intracerebral Hemorrhage Evacuation (MISTIE-III) trial; a multicenter, randomized phase III trial conducted between 2013 and 2018.^4^ MISTIE-III enrolled adults with spontaneous ICH over 30 ml in volume. Participants were randomized to standard medical management, as per local guidelines, or MIS and alteplase, using a cerebral catheter. Figure 1 details the inclusion of participants in this secondary analysis.

**Figure 1:**
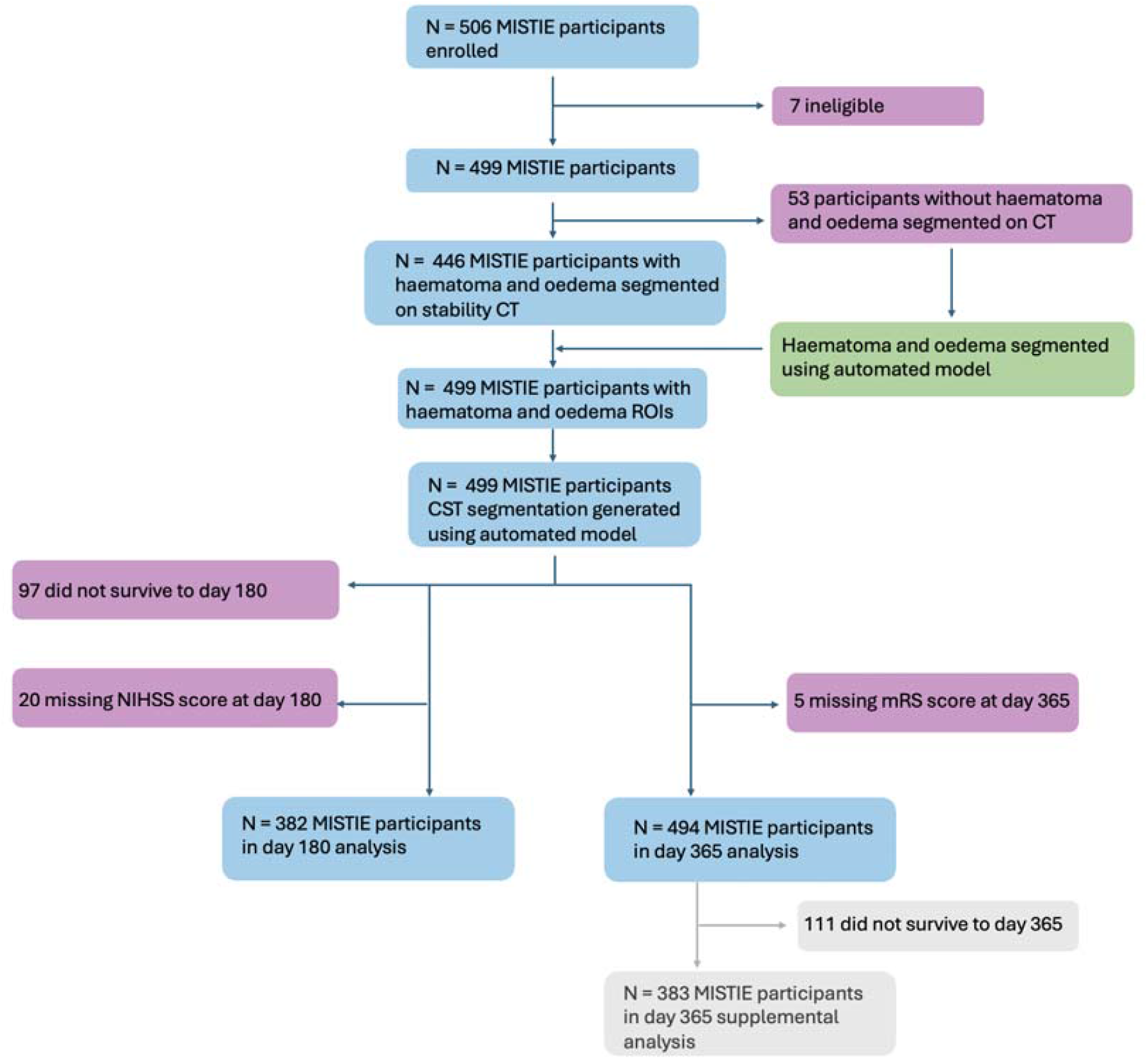
Inclusion flow diagram detailing the participants included in each statistical analysis. Patients missing data at any timepoint, due to death and/or loss-to-follow up, were not included in the analysis of that timepoint

#### Ethics

Our secondary analysis was conducted within the original data use agreements^4^ and no additional ethical approval was required.

#### Imaging

The MISTIE-III protocol required a stability CT scan ≥ 6 hours after the diagnostic CT. If ICH volume had increased by > 5 ml, further CTs were performed every 12 hours until the hematoma was no longer increasing in volume. The final stability CT scan (N=499) was used for our analyses to ensure we accounted for the final hematoma volume.

#### Clinical measurements

We used the motor National Institutes of Health Stroke Scale (NIHSS) at day 180 as the primary outcome as it is used in routine clinical care and typically reaches a plateau by six months^18^. The motor component of the NIHSS score is the sum of the arm, face and leg scores (questions 4, 5 and 6), ranging from 0 to 19.^17^ We chose the modified Rankin Scale (mRS) at day 365 as a secondary outcome to reflect the primary outcome of MISTIE-III^4^.

### Hematoma and edema segmentation

#### Manual segmentation

Hematoma and PHE have been previously segmented for the stability CT scan for 446 participants using a validated, semi-automated approach.^19^ Segmentation was conducted by two experienced staff members at the BIOS Imaging Center (Johns Hopkins University, USA) using OsiriX imaging software (OsiriX v. 4.1, Pixmeo; Geneva, Switzerland).

#### Automated imputation

53 participants were missing hematoma and PHE segmentations on the final stability CT scan. An automated nnU-Net model was trained on the 446 participants with manual segmentations (training N=365, testing N=90), over 1000 epochs using a combination Dice and Cross Entropy loss on two NVIDIA a100 GPUs^20^. The model was ensembled over 5 folds, and performance on the testing dataset was assessed with the Dice similarity coefficient (DSC). Hematoma and PHE segmentations were then generated for the 53 stability CT scans missing hematoma and PHE segmentations.

### Corticospinal tract risk score

We previously developed an automated method for segmenting the CST on non-contrast CT using a dataset of patients with ICH who underwent both diagnostic CT and diffusion tensor MRI within a short timeframe to train a deep learning-based CST segmentation model. ^16^ The nnU-Net based model produced segmentations of the CST from diagnostic CT scans alone, achieving a Dice similarity coefficient of 57% when compared with CST labels derived from diffusion tensor tractography.

This CST segmentation model was used to derive a CST risk score reflecting increasing risk of CST damage. CST risk was classified from the interaction of the CST with the hematoma and PHE. Participants were classified to one of four levels, reflecting an increasing theoretical risk of CST injury and thus a greater risk of poor motor outcome:

- **CST risk 0** (No risk): There is no overlap between the CST and the PHE or hematoma.
- **CST risk 1** (PHE infiltration): There is some overlap between the CST and the PHE. The hematoma does not infiltrate the CST.
- **CST risk 2** (Hematoma infiltration): There is some overlap between the CST and the hematoma.
- **CST risk 3** (CST interruption): The CST is not continuous; the hematoma has enveloped the CST.

When multiple risk categories were present, the highest category was used. CST risk 3 was defined as discontinuity in the predicted CST segmentation in the axial plane due to complete envelopment by the hematoma.

### Statistical analysis

#### Primary analysis

Primary analysis investigated whether CST risk modified the effect of MIS on motor NIHSS at day 180 using multivariable linear regression. The model included CST risk, age, treatment group (surgical vs. medical), the log-transformed ICH volume, IVH volume and baseline motor NIHSS. An interaction term between CST risk score and treatment group was included to test whether CST risk score modified the effect of surgical intervention on outcome. CST risk 3 was used as the reference category.

#### Secondary analysis

To reflect the primary endpoint of the MISTIE-III trial, secondary analysis investigated whether CST risk score modified the effect of MIS on mRS score at day 365 using multivariable ordinal regression. Covariates were the same as for the primary analysis, except baseline Glasgow Coma Scale score replaced baseline motor NIHSS. Due to small sample size in the mRS 0 category violating the proportional odds assumption, mRS categories 0 and 1 were collapsed. This analysis was repeated with mRS at day 180.

The primary analysis was repeated with motor NIHSS day 365 as the dependent variable to investigate the effects of CST risk score on long term motor outcome after surgery.

#### Sensitivity analysis

To assess whether associations with mRS were driven by mortality, the ordinal regression was restricted to patients who survived to day 365 (mRS 0-5, excluding mRS 6).

As MISTIE-III found functional outcome was improved when an end of treatment ICH volume of >15ml was achieved, the primary and mRS day 365 analysis repeated, restricting the surgical group to end of treatment ICH volume >15ml.

## Results

### Baseline characteristics

Baseline characteristics of all MISTIE-III participants by CST risk group are shown in Table 1. Participants with higher levels of CST risk tended to have lower GCS and higher NIHSS scores but ICH volumes were comparable. Deep ICH was more common in the higher risk CST groups and all but one of the no risk groups were lobar ICH.

**Table 1:** Baseline demographics of the MISTIE-III clinical trial, split by CST risk.

| **Variable** | **Total [506]** | **No risk [37]** | **PHE**<br>**infiltration**<br>**[187]** | **Hematoma**<br>**infiltration [120]** | **Tract**<br>**interruption**<br>**[162]** |
| --- | --- | --- | --- | --- | --- |
| **Surgery**[N] | 255 (51%) | 16 (44%) | 86 (46%) | 65 (55%) | 88 (55%) |
| **Medical**[N] | 251 (50%) | 21 (57%) | 101 (54%) | 55 (46%) | 74 (46%) |
| **Location**[N] |  |  |  |  |  |
| Deep | 307 (61%) | 1 (3%) | 80 (43%) | 90 (75%) | 136 (84%) |
| Lobar | 192 (38%) | 36 (98%) | 101 (54%) | 30 (25%) | 25 (16%) |
| **Age** mean(SD) | 61.2±12.3 | 67.6 ± 10.1 | 62.6 ± 12.2 | 58 ± 12.7 | 60.7 ± 11.9 |
| **Age**[range] | [28–90] | [40–83] | [29–90] | [28–88] | [31–85] |
| **Race**[N] |  |  |  |  |  |
| White | 379 (75%) | 32 (87%) | 149 (80%) | 82 (69%) | 116 (72%) |
| African-American | 89 (18%) | 4 (11%) | 29 (16%) | 26 (22%) | 30 (19%) |
| Asian | 30 (6%) | 1 (3%) | 8 (5%) | 9 (8%) | 12 (8%) |
| Hawaiian or Pacific | 3 (1%) | 0 (0%) | 0 (0%) | 0 (0%) | 3 (2%) |
| Native American | 2 (1%) | 0 (0%) | 0 (0%) | 1 (1%) | 1 (1%) |
| **GCS Pre-Rand** median[IQR] | 10 [8–13] | 13 [10–13] | 11[9–13] | 10 [8–13] | 9 [8–12] |
| **GCS Pre-Rand**[range] | [3–15] | [8–15] | [3–15] | [3–15] | [5–15] |
| Mild GCS[N] | 93 (19%) | 9 (25%) | 42 (23%) | 26 (22%) | 16 (10%) |
| Moderate GCS[N] | 284 (57%) | 27 (73%) | 110 (59%) | 59 (50%) | 88 (55%) |
| Severe GCS[N] | 129 (26%) | 1 (3%) | 35 (19%) | 35 (30%) | 58 (36%) |
| **NIHSS Pre-Rand** median[IQR] | 19 [15–23] | 13 [7–16] | 18 [14.5–22] | 19 [15–24.3] | 20.5 [18–24] |
| **NIHSS Pre-Rand**[range] | [1–40] | [1–29] | [2 – 40] | [4–39] | [6–40] |
| Minor NIHSS[N] | 9 (2%) | 4 (11%) | 4 (3%) | 1 (1%) | 0 (0%) |
| Moderate NIHSS[N] | 127 (26%) | 22 (60%) | 56 (30%) | 30 (25%) | 19 (12%) |
| Moderate-Severe NIHSS[N] | 167 (33%) | 6 (17%) | 59 (32%) | 40 (34%) | 62 (39%) |
| Severe NIHSS[N] | 203 (41%) | 5 (14%) | 68 (37%) | 49 (41%) | 81 (50%) |
| **Motor NIHSS** median[IQR] | 10 [8–12] | 4 [2–8] | 9 [7–11] | 10 [8–13] | 10 [9–12] |
| **Motor NIHSS**[range] | [0–19] | [0–14] | [0–19] | [1–19] | [1–19] |
| **ICH Volume** median[IQR] | 45.3 [35.5–58.5] | 41.4 [33.1–48.8] | 44.2 [35–57.1] | 40.7 [33.1–53.8] | 50.7 [39.5–67.6] |
| **ICH Volume**[range] | [20.9–127.1] | [24.5–105.8] | [20.9–118.8] | [24.4–80.4] | [21.7–127.1] |
| **IVH**[N] | 316 (62%) | 24 (65%) | 101 (54%) | 74 (61%) | 117 (72%) |
| **IVH Volume** mean (SD)(mL) | 2.7 ± 5.7 | 1.5 ± 2.2 | 2 ± 4.3 | 2.4 ± 5.1 | 4 ± 7.5 |
| **IVH Volume**[range] | [0–61.8] | [0–8.2] | [0–28.8] | [0–29.5] | [0–61.8] |

### Missing segmentation model performance

The mean DSC between ground truth segmentations and the model predictions for the test set (n=90) were 0.95 and 0.80 for hematoma and PHE, indicating sufficient accuracy.

### CST segmentations

CST segmentations for four MISTIE-III participants are shown in Figure 2.

**Figure 2:**
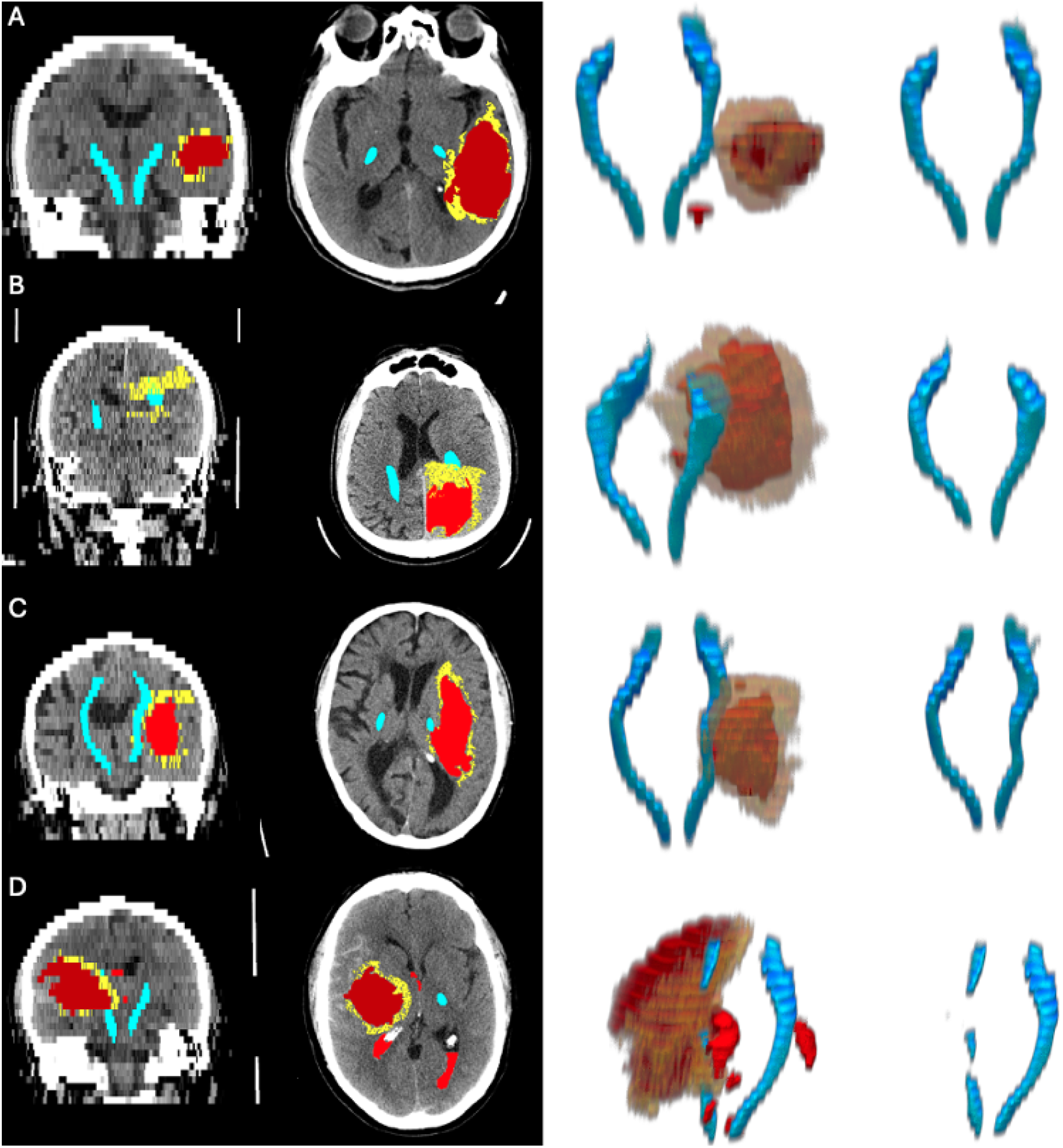
Four MISTIE-III participants representing each of the four levels of CST risk. CST segmentations are in blue, hematoma segmentations in red, and PHE segmentations in yellow. On the left are 2D slices from stability CT, and on the right are 3D reconstructions of the CST, hematoma and PHE. **A**: No CST risk, **B**: PHE infiltration, **C**: Hematoma infiltration, **D**: Tract interruption

### Association between surgical treatment group, corticospinal tract risk and outcome after stroke

The primary multivariable linear regression model with motor NIHSS score at day 180 as outcome is shown in Table 2 and Figure 3. Compared to the reference category of CST interruption, no risk and PHE infiltration were significantly associated with lower motor NIHSS. The interaction term between CST risk score and treatment group was significant, with hematoma infiltration (CST risk score 2) significantly associated with less severe motor NIHSS score at day 180 in the surgery group. An interaction plot depicts the relationship between motor NIHSS score at day 180 and treatment group for different CST risk scores in Figure 3.

**Table 2:** Results of the multivariate linear regression analysis for motor NIHSS at day 180 and the ordinal regression analysis for mRS at day 365.

| Variable | Motor NIHSS day 180 $\beta$<br>(95% CI) | p-value | mRS (ordinal) OR (95%<br>CI) | p-value |
| --- | --- | --- | --- | --- |
| **Treatment group[surgery]** | 0.97 (-0.2-2.1) | 0.10 | 0.72 (0.41-1.27) | 0.257 |
| **No risk** | -3.77 (-5.8--1.7) | 0.0003 | 0.27 (0.10-0.74) | 0.011 |
| **PHE infiltration** | -2.30 (-3.5--1.1) | 0.0002 | 0.41 (0.23-0.74) | 0.003 |
| **Hematoma infiltration** | -0.42 (-1.7-0.9) | 0.52 | 0.59 (0.31-1.13) | 0.110 |
| **Tract interruption** | ref | ref | ref | ref |
| **Baseline motor NIHSS** | 0.40 (0.3-0.5) | <0.0001 | — | — |
| **GCS at randomization** | — | — | 0.83 (0.77-0.89) | <0.0001 |
| **Age (years)** | 0.04 (0.0-0.1) | 0.003 | 1.06 (1.04-1.07) | <0.0001 |
| **IVH volume** | 0.33 (-0.7-1.3) | 0.51 | 1.03 (1.00-1.06) | 0.046 |
| **log(ICH volume)** | -0.01 (-0.1-0.1) | 0.79 | 3.74 (2.25-6.23) | <0.0001 |
| **Treatment group [surgery] × No risk** | 0.64 (-2-3.3) | 0.64 | 0.23 (0.05-0.97) | 0.045 |
| **Treatment group [surgery] × PHE infiltration** | -0.73 (-2.3-0.9) | 0.35 | 1.34 (0.61-2.92) | 0.471 |
| **Treatment group [surgery] × Hematoma infiltration** | -2.07 (-3.8--0.4) | 0.016 | 1.34 (0.57-3.16) | 0.505 |

**Figure 3:**
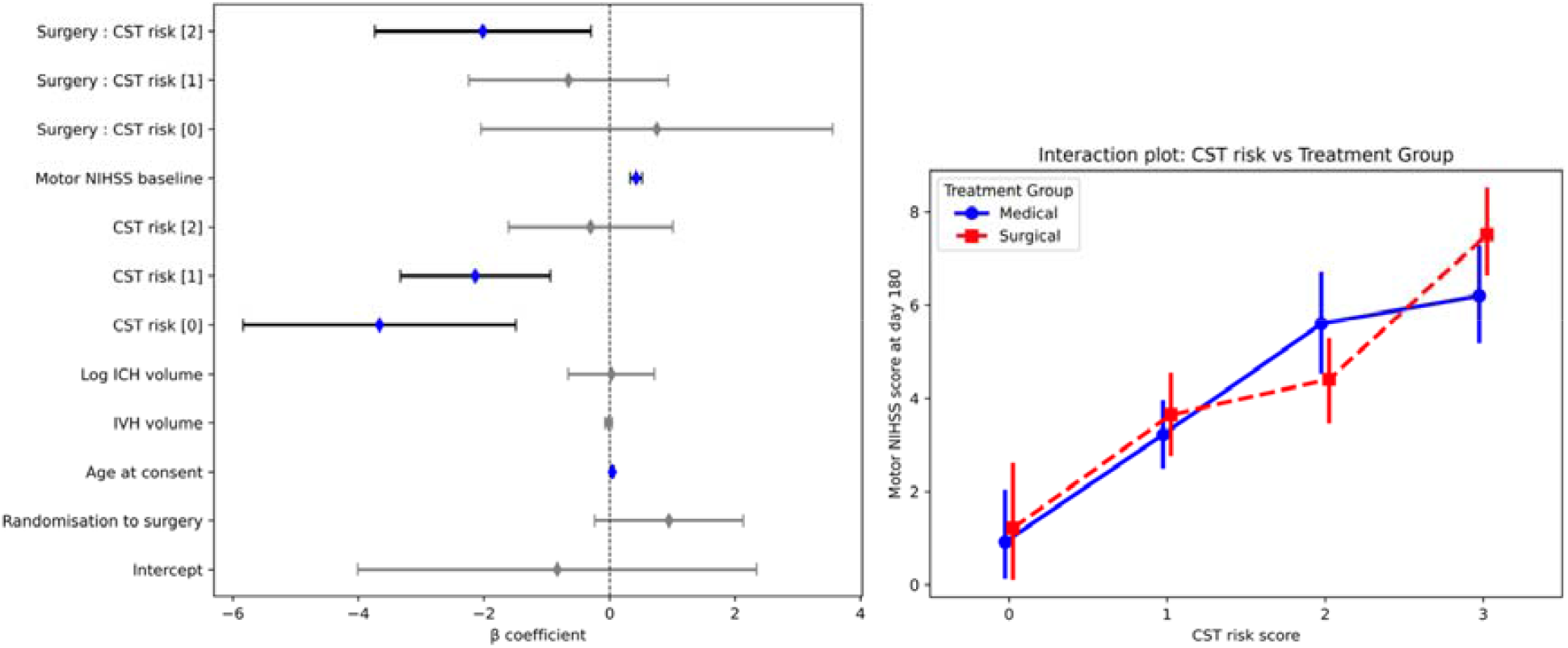
Left: Forest plot showing -coefficients of the multivariable linear regression analysis for motor NIHSS at day 180 (statistically significant variables (p<0.05) in blue). Right: An interaction plot showing the relationship between treatment group and unadjusted motor NIHSS at day 180 for different CST risk scores (0=no risk, 1=PHE infiltration, 2=hematoma infiltration, 3=CST interruption)

Secondary analyses broadly supported these results. Similar associations were observed for motor NIHSS day 365 (Supplemental Table 1), including the significant interaction term between hematoma infiltration and surgical intervention.

Secondary ordinal regression analysis testing the association between surgery and CST risk with mRS at day 365 met the proportional odds assumption (*p* for proportional odds = 0.42) and is shown in Table 2. No CST risk and PHE infiltration were associated with lower odds of an unfavorable outcome. The interaction term was significant; with no CST risk significantly associated with less severe mRS score at day 365 in the surgery group. When repeated for mRS day 180, similar associations were observed, however the interaction term between no CST risk and surgery did not reach significance (Supplemental table 1).

When the primary analysis was restricted to surgical patients for which an end of treatment hematoma volume of <15ml was achieved, similar associations with motor NIHSS at day 180 were observed, however the interaction term between no CST risk and surgery was no longer significantly associated with improved mRS at day 365 (Supplemental table 2).

When the secondary mRS analyses was repeated in patients who survived to day 365, results were broadly similar (Supplemental Table 3).

## Discussion

In our secondary analysis of the MISTIE-III trial, we have demonstrated a progressively increasing risk of poor motor outcome and worse mRS with increasing risk of CST injury along our risk classification, suggesting that it may serve as a useful independent prognostic factor for motor outcome and disability. Furthermore, CST risk is a significant modifier of the effect of the MISTIE-III surgical intervention on motor outcome at day 180, with surgery reducing day 180 motor NIHSS scores by two points more in participants with hematoma infiltration, compared to participants with CST interruption. The significant interaction term between surgery and hematoma infiltration suggests patients with CST infiltration may benefit most from MIS but given the exploratory and retrospective nature of our study, this requires further investigation.

A significant interaction between no CST risk and surgery was seen in our model with mRS at day 365 as outcome. This suggests that whilst surgical evacuation of hematomas infiltrating the CST results in improved motor NIHSS, overall disability is reduced by surgical hematoma evacuation in patients that have no risk of CST damage. This is to be interpreted cautiously, as the confidence interval is wide, and the sample size of patients with no CST risk who received surgery is relatively small. This significance disappears when we restricted the analysis to surgical patients in which end of treatment hematoma volumes less than 15ml were achieved, whereas the interaction term in the NIHSS analysis remained significant, further suggesting the interaction between no CST risk and surgery may be due to small sample size. Whilst motor function is a major determinant of mRS, other domains of neurological deficit and other factors not captured by the mRS will also contribute, potentially weakening the association between CST injury and mRS when compared to NIHSS at day 180.

In hyperacute ICH, the CST may be compressed by the hematoma, resulting in ischemia, physical disruption and reduced axonal conduction.^14,15^ After the initial insult, secondary injury may occur, resulting from the release of heme, iron and free radicals and the deleterious inflammatory response in the perihematomal brain.^21^ Surgical evacuation of the hematoma theoretically mitigates the effects of both mass effect and secondary injury, which would result in further damage in the days after stroke^14^. The MISTIE procedure included the administration of alteplase. In animal models of ischemic stroke, alteplase has been shown to have nonthrombolytic effects^22^ which may increase CST injury, warranting further study in trials of MIS without alteplase. MISTIE-III and MIND commenced MIS within approximately 72 hours of symptom onset and were neutral, whereas ENRICH tested MIS within 24 hours and was positive, suggesting that timing may also play an important role.^4–6^

When there is no risk to the CST prior to surgery, it follows that surgery cannot alter motor outcome, as observed. However, we see a reduction in overall disability in participants in the no risk group who received surgery, suggesting beneficial effects on other neurological deficits and/or a reduction in the risk of death, though this should be interpreted cautiously due to small sample size. For participants with PHE infiltration, we observe no interaction with surgery. It is possible that the reduction in risk to the CST from MIS may not outweigh the risks of MIS, or that the use of alteplase offsets any benefit. Surgery in participants with hematoma infiltration of the CST was significantly associated with better motor NIHSS at day 180, which is consistent with the hypothesis that MIS may prevent further damage to the CST, though this does not translate to significant heterogeneity in the mRS model. When the CST is interrupted by the hematoma, MIS may be futile and may even worsen motor NIHSS at day 180, as suggested by inspection of the unadjusted data in the interaction plot in Figure 3.

Our results suggest that a more nuanced approach to location in relation to the CST may allow identification of a subgroup of deep ICHs who will benefit from MIS. This has been highlighted by the recent clinical trial landscape: the broad inclusion criteria of the MIND trial which included 2/3 of patients with deep haemorrhage yielded an overall negative result^6^, whereas the positive findings of the ENRICH trial were heavily dependent on restricting interventions to lobar haemorrhages^5^. Because a substantial proportion of real-world patients present with deep ICH, abandoning surgical options for this entire cohort based on broad anatomical location alone may be premature. The CST risk score differentiates between patients with haematoma infiltration of the CST (33% lobar) and patients with full CST interruption by the haematoma (18% lobar). Both groups both had a high proportion of deep haematomas but different outcomes and different responses to surgical haematoma evacuation. Prospective trials are needed to test this hypothesis, and our method could be easily included in an acute stroke pipeline to identify eligible participants.

Our study benefits from the robust and rich dataset collected as part of the MISTIE-III trial. We were able to use stability scans rather than baseline scans, which allows us to include any hematoma expansion after the diagnostic CT. The great majority of the scans we used had accurate and robust manual segmentations of hematoma and PHE conducted by an experienced clinical trial imaging center using a validated method. The pipeline we present for assessing CST risk is automated and uses diagnostic CT scans alone. This makes our method quick and accessible to stroke centers without advanced MRI imaging, such as diffusion tensor imaging, or transcranial magnetic stimulation in their stroke pipeline.

Our study has several limitations. Firstly, the CST segmentation model reproduced DTI-tractography-like segmentations from CT scans alone with a DSC of 57%^16^, which though more consistent than manual CT-based approaches, loses some information contained within gold standard DTI tractography. Accordingly, the CST risk scale should be interpreted as an imaging-derived approximation of CST injury and may not reflect the true level of axonal damage. Structural integrity is also only one aspect of CST damage; functional integrity, measured using transcranial magnetic stimulation, has been shown to have a distinct relationship with motor outcome after stroke^7–9^. Secondly, the outcome measure chosen was the motor component of the NIHSS score. A comprehensive motor score, such as the Fugl-Meyer score, would likely be more sensitive to CST integrity. Finally, we have measured PHE on CT at a single and variable time point. As PHE evolves over the acute phase, differences in scan time may mean the PHE is measured at varying stages in its evolution^23,24^ potentially adding further heterogeneity to the assessment of secondary CST injury risk.

In conclusion, the CST risk score is strongly and independently associated with long term functional outcome after stroke. Level of CST risk modified the effect of surgery on motor outcome 180 days after ICH, presenting the hypothesis that patients with hematoma infiltration of the CST may benefit from MIS. The CST risk score, which can be automatically computed from widely available diagnostic imaging, could be explored as a new biomarker for future surgical trials.

## Acknowledgements

We would like to thank the participants of MISTIE-III and their families, and the MISTIE-III investigators. We also acknowledge support from the Natalie Kate Moss Trust.

## Statements and Declarations

### Ethical considerations and consent

Each hospital included in MISTIE-III obtained local institutional review board or ethics committee approval, described in the study protocol. Informed consent was obtained from all patients or legal representatives according to the MISTIE-III study protocol. No identifiable individual data has been included.

### Data availability

This is a secondary analysis of the MISTIE-III trial. Data can be accessed through NINDS or by contacting the MISTIE-III investigators

## Supplemental materials

Supplemental tables 1-3

