## Supplemental 1 for "Corticospinal tract risk modifies motor recovery after minimally invasive surgery for intracerebral hemorrhage: a secondary analysis of MISTIE-III"

**Supplemental Table 1**: Motor *NIHSS day 365 multivariate linear regression and mRS day 180 multivariate ordinal regression model results*

A table displaying the multivariate linear regression for the association between CST risk, treatment, and motor NIHSS at day 365; and the multivariate ordinal regression analysis investigating the association between CST risk combined with surgical treatment, and motor mRS at day 180.

| **Variable** | **Motor NIHSS day 365 β (95% CI)** | **p-value** | **mRS (ordinal) OR (95% CI)** | **p-value** |
| --- | --- | --- | --- | --- |
| **Treatment group [surgery]** | 1.34(0.1−2.56) | 0.03 | 0.73 (0.41–1.22) | 0.26 |
| **No risk** | -3.08(-5.2−-1.0) | 0.004 | 0.27 (0.10–1.35) | 0.012 |
| **PHE infiltration** | -1.47(-2.69−-0.25) | 0.02 | 0.41 (0.22–0.74) | 0.003 |
| **Hematoma infiltration** | -0.31(-1.0−1.7) | 0.65 | 0.58 (0.30–1.11) | 0.098 |
| **Tract interruption** | ref | ref | ref | ref |
| **Sum of NIHSS motor domains (4–6) at baseline** | 0.38(0.3−0.5) | <0.0001 | — | — |
| **GCS at randomisation** | — | — | 0.83 (0.74–0.90) | <0.0001 |
| **Age (years)** | 0.04(0.0−0.1) | 0.002 | 1.06 (1.04–1.07) | <0.0001 |
| **IVH volume** | -0.03(-0.1−0.0) | 0.44 | 1.03 (1.00–1.06) | 0.045 |
| **log(ICH volume)** | 0.12(-0.9−1.1) | 0.82 | 3.67 (2.23–6.05) | <0.0001 |
| **Treatment group [surgery] × No risk** | -0.12(-2.8−2.6) | 0.93 | 0.26 (0.06–1.06) | 0.061 |
| **Treatment group [surgery] × PHE infiltration** | -1.28(-2.9−0.3) | 0.12 | 1.35 (0.62–2.97) | 0.45 |
| **Treatment group [surgery] × Hematoma infiltration** | -2.14(-3.9−-0.4) | 0.018 | 1.38 (0.58–5.75) | 0.47 |

**Supplemental Table 2**: Motor *NIHSS day 180 multivariate linear regression and mRS day 365 multivariate ordinal regression model results, restricted to patients in the medical group or in the surgical group who achieved <15ml clot reduction.*

A table displaying the multivariate linear regression for the association between CST risk, treatment, and motor NIHSS at day 365; and the multivariate ordinal regression analysis investigating the association between CST risk combined with surgical treatment, and motor mRS at day 180.

| **Variable** | **Motor NIHSS day 180 β (95% CI)** | **p-value** | **mRS (ordinal) OR (95% CI)** | **p-value** |
| --- | --- | --- | --- | --- |
| **Treatment group [surgery]** | 0.61(0.72 −1.9) | 0.37 | 0.57 (0.30–1.11) | 0.12 |
| **No risk** | -3.7(-5.7−-1.7) | 0.0003 | 0.29 (0.10–0.82) | 0.015 |
| **PHE infiltration** | -2.27(-3.4−-1.1) | 0.0001 | 0.43 (0.24–0.77) | 0.005 |
| **Hematoma infiltration** | -0.44(-1.7−0.8) | 0.49 | 0.59 (0.30–1.13) | 0.11 |
| **Tract interruption** | ref | ref | ref | ref |
| **Sum of NIHSS motor domains (4–6) at baseline** | 0.42(0.3−0.5) | <0.0001 | — | — |
| **GCS at randomisation** | — | — | 0.82 (0.06–0.90) | <0.0001 |
| **Age (years)** | 0.03(0.0−0.1) | 0.03 | 1.05 (1.04–1.07) | <0.0001 |
| **IVH volume** | -0.05(-0.1−0.0) | 0.22 | 1.02 (0.98–1.06) | 0.39 |
| **log(ICH volume)** | 0.28(-0.9−1.5) | 0.65 | 3.29 (1.75–6.05) | <0.0001 |
| **Treatment group [surgery] × No risk** | 0.19(-2.8−3.2) | 0.9 | 0.51 (0.10–2.61) | 0.42 |
| **Treatment group [surgery] × PHE infiltration** | -0.81(-2.6−0.9) | 0.37 | 1.15 (0.45–2.92) | 0.77 |
| **Treatment group [surgery] × Hematoma infiltration** | -2.18(-4−-0.4) | 0.019 | 1.63 (0.61–4.31) | 0.33 |

**Supplemental Table 3**: mRS *multivariate ordinal regression*

The results of the multivariate ordinal regression (proportional odds) analysis investigating the association between CST risk combined with surgical treatment, and outcome, as assessed by motor mRS (excluding mRS 6) at day 365.

| **Variable** | **mRS (ordinal) OR (95% CI)** | **p-value** |
| --- | --- | --- |
| **Treatment group [surgery]** | 0.76 (0.43–1.33) | 0.333 |
| **No risk** | 0.28 (0.10–0.78) | 0.014 |
| **Edema infiltration** | 0.43 (0.24–0.79) | 0.006 |
| **Hematoma infiltration** | 0.60 (0.31–1.16) | 0.127 |
| **Tract interruption** | ref | ref |
| **Age (years)** | 1.06 (1.04–1.07) | <0.001 |
| **GCS at randomisation** | 0.84 (0.78–0.90) | <0.001 |
| **log(ICH volume)** | 3.76 (2.24–6.31) | <0.001 |
| **IVH volume** | 1.03 (1.00–1.06) | 0.044 |
| **Treatment group [surgery] × No risk** | 0.22 (0.05–0.93) | 0.039 |
| **Treatment group [surgery] × Edema infiltration** | 1.28 (0.58–2.84) | 0.542 |
| **Treatment group [surgery] × Hematoma infiltration** | 1.33 (0.56–3.15) | 0.522 |
